# Effect of Single versus Multiple Prophylactic Antibiotic Doses on Revision Rates due to Infection after Total Hip Arthroplasty: Protocol for A Target Trial Emulation Study Using Danish National Registries

**DOI:** 10.1101/2025.04.13.25325741

**Authors:** Armita Armina Abedi, Marie Anneberg Brahe, Alma Becic Pedersen, Claus Varnum, Robin Christensen, Søren Overgaard

**Author notes:** **Contact details for further information:** Armita Armina Abedi. **Statistical Analyst:** Armita Armina Abedi.

## Abstract

**Introduction:** Prophylactic antibiotics can reduce the risk of prosthetic joint infection (PJI), which is among the most severe complications following total hip arthroplasty (THA). Despite its importance, there is no consensus on the duration of prophylactic antibiotics, and current recommendations describe the use of either one single dose or up to 24 hours with multiple doses as options for antibiotic prophylaxis.

**Objective:** To compare a single dose versus multiple doses of prophylactic antibiotics administered within 24 hours, on the risk of revision due to infection within the first 90 days after THA. Noninferiority will be demonstrated if the upper limit of the two-sided 95% confidence interval for the odds ratio is less than 1.4 for the single dose compared with multiple doses of antibiotic prophylaxis administered within 24 hours of the index surgery.

**Design:** An emulated target trial combining data from the Danish Hip Arthroplasty Register and the Danish Microbiology Database naively assuming that operated patients were randomized to either single dose or multiple doses of prophylactic antibiotics administered within 24 hours.

**Setting:** All departments of orthopedic surgery in Denmark, from January 1^st^, 2010, to December 31^st^, 2020.

**Participants:** Patients with age ≥18 years, undergoing primary THA for all reasons except due to acute or sequelae from proximal femoral or pelvic fractures or bone tumor or metastasis will be included.

**Intervention:** A single dose of preoperatively administered prophylactic antibiotic administered preoperatively.

**Comparator:** Multiple doses of prophylactic antibiotics administered within 24 hours of THA.

**Outcomes:** The primary outcome is the revision rates due to infection within 90 days after primary THA. Secondary outcomes within the first 90 days post-surgery include (*i*) risk of revision due to infection after primary THA (*i*) any revision after THA (*ii*) risk of potential PJI (*iii*) hospital-treated infections, (i*v*) community-based antibiotic use, and (*v*) mortality rate and furthermore, within 365 days post-surgery (*vi*) risk of revision due to infection after primary. All outcome measures will be extracted from national databases.

**Analyses:** Analyses will be based on the Intention to Treat (ITT) population (i.e., all patients having a THA where some dose regimen of antibiotics is registered). The main analyses will explicitly emulate the components of the target trial protocol using the observational data. Based on the eligible individuals, assigned to a treatment strategy based on their data, following them from treatment assignment (time zero) until outcome or the end of follow-up (90 and 365 days from surgery, respectviely), and conducting the same analysis as the target trial. However, we will adjust for baseline confounders to approximate random treatment assignment and balance baseline demographic and clinical characteristics (i.e., pre-surgery covariates), we will use stabilized inverse probability treatment weighting (sIPTW).

**Motivation:** We believe that the results of this trial will deliver important knowledge on optimal antibiotic prophylaxis dosage practice for primary THA.

## BACKGROUND

The most feared complication of THA is prosthetic joint infection (PJI), and the incidence seems to be increasing depending on definition and capture methods. In Denmark, the incidence of PJI varies from 0.5% to up to around 3% on department level (1) and is associated with increased morbidity and mortality (2–5). Perioperative antimicrobial prophylaxis is a well-established and documented part of standard care to reduce the risk of PJI (6–10). However, there exists no consensus on the duration of antibiotic prophylaxis. Some associations advocate for using one single preoperative dose (11, 12) and other guidelines recommend antibiotic prophylaxis for up to 24 hours from index surgery (13–15). The Danish national guideline advises the administration of antibiotic prophylaxis using either cloxacillin or the second-generation cephalosporin, cefuroxime with discontinuation within 24 hours after surgery (16). The choice of antibiotic and duration vary among the different orthopedic departments in Denmark.

The possibility of reducing the use of post-operative antibiotics without compromising patient safety may pose multiple advantages for the patient and for society. The use of antibiotics is regarded as the primary driver of the worldwide antimicrobial resistance crisis making treatment of common infections difficult or even impossible (17–19). Based on systematic reviews, both single-dose and multiple-dose antibiotic prophylaxis regimens show comparable results in the prevention of infection after THA (20–22). A large retrospective study (23) suggests that a single dose may be non-inferior to multiple doses of prophylactic antibiotics in the prevention of PJI after THA. Furthermore, a comparable risk of complete revision for infection between a single-dose versus multiple-dose antibiotic prophylaxes was found in a recent large observational register-based study of the Dutch Arthroplasty Register (24). The study collected data on antibiotic prophylaxis regimens through a national audit as individual patient-level data was unavailable (25) and the outcome, and revision for infection were solely captured through the Dutch Arthroplasty register. Most national registries are not designed to capture infection and thus underreporting of infection may be present (26, 27).

Combining data from the Danish Hip Arthroplasty Register (DHR) with the Danish Microbiology Database (MiBa) has been shown to significantly improve the diagnosis of PJI (27). A validated algorithm using data from DHR and MiBa, using 2 or more identical positive bacterial cultures, increased the positive predictive value (PPV) from 77% to 98 % and the negative predictive value (NPV) from 92% to 98% compared to the registration alone by the surgeon in DHR (28). By applying this well-validated method for the capture of PJI, we aim to evaluate the risk of revision due to infection between single doses versus multiple doses of antibiotics more accurately. An observational study with real-world data may add valuable knowledge on antibiotic prophylaxis duration and the risk of PJI and a large dataset may allow for an adequate power in the detection of rare events such as PJI (29). However, the outcomes from an observational study may be prone to bias due to a lack of randomization and stringent research protocol (30). By applying an emulated target trial (ETT), observational data may be used to mimic a randomized controlled trial (RCT) as closely as possible (30, 31).

This study aims to evaluate the effect of a single versus multiple doses of antibiotic prophylaxis administered within 24 hours from index surgery, on the risk of revision due to infection following the first 90 days after primary THA. We hypothesize that a single dose of prophylactic antibiotic is non-inferior to multiple doses of prophylactic antibiotics.

### Objectives

Our primary effectiveness objective is to compare the effect of a single versus multiple doses of antibiotic prophylaxis administered within 24 hours from index surgery, on the risk of revision due to infection following the first 90 days after primary THA. Noninferiority will be demonstrated if the upper limit of the two-sided 95% confidence interval for the odds ratio is less than 1.4 for the single dose compared with multiple doses of antibiotic prophylaxis administered within 24 hours of the index surgery.

Secondary effectiveness objectives, we will also assess further outcomes collected up to 90 days from index surgery, any revision after THA, potential PJI referred to as *PJI-likely*, hospital-treated infections (Not PJI or PJI-likely), community-based antibiotic use and mortality as well as the risk of revision due to infection up to 365 days from index surgery.

## METHODS

### Study Design and Setting

The study protocol was designed following the ETT framework (29, 30), and an emulation of this target trial is summarized in Appendix A, Table 1. Data will be reported according to the RECORD guidelines (32) and stored on Statistics Denmark’s servers.

All patients receiving a first-time primary THA between January 1^st^, 2010, to December 31^st^, 2020, will be included corresponding to approximately 10,000 hips included per year corresponding to approximately 110,000 hips during the study period. All procedures will be identified from the DHR and inclusion up to 31.12.2020 enables follow-up until 31.12.2021 allowing for at least a 1-year follow-up for all patients. The type and duration of prophylactic antibiotics are registered for each procedure reported to DHR. Patients operated with each antibiotic prophylaxis duration will be divided into 2 groups as reported to DHR:

– one single preoperative dose
– up to 24 hours of prophylaxis

Cases with missing registration for antibiotic prophylaxis duration will be excluded as well as durations for more than 24 hours of prophylaxis. Revisions performed within the first 90 days and 365 days after primary surgery are included and separated by cause of revision. Information on patient characteristics including comorbidities and demographic data will be collected through several databases further elaborated in the section on *Data Sources*.

### Eligibility criteria

All patients age ≥ 18 years receiving a primary THA

➢ Only the first operated hip during the study period will be included

Exclusion criteria

➢ Patients receiving a primary THA due to either acute or sequelae from proximal femoral or pelvic fractures
➢ Patients receiving a primary THA due to bone tumor or metastasis

#### Treatment Strategies

Antibiotic prophylaxis with a single dose will be compared with multiple doses given within 24 hours after primary THA as reported in DHR by the surgeon. The eligible patients will have received either the penicillin dicloxacillin or the second-generation cephalosporin cefuroxime. In the event of a cephalosporin or general beta-lactam allergy, clindamycin or another appropriate antibiotic may have been utilized, and these cases will also be included. Postoperative management will be the same for all patients in the two antibiotic prophylaxis duration groups. Discontinuation of the assigned strategy could be due to an allergic reaction to the antibiotic treatment.

#### Treatment Assignment

Ideally, in an RCT setting, the patients should be randomly allocated to either a single preoperative dose of antibiotic prophylaxis or continued prophylaxis for up to 24 hours. However, due to the observational character of the data collected for this study, randomization will not be possible. For the ETT, treatment assignment will be based on the antibiotic treatment duration registered in DHR by the surgeon. The assignment will be based on routine clinical practice following standard departmental instructions for antibiotic prophylaxis for primary THA at the given period or chosen by the surgeon based on individual clinical assessment. Eligible patients will be divided into one of the following two groups of antibiotic prophylaxis duration as reported to DHR: one single preoperative dose and up to 24 hours of prophylaxis after primary THA. The propensity of the treatment assignment to the single-dose group or the multiple-dose group (within 24 hours) will be based on identified predictors of treatment assignment and the analysis will be weighted for this propensity (32).

#### Time Zero and Follow-up

Time zero is defined as the date of primary THA, at which the patient receives the first dose of antibiotic prophylaxis. The type and duration of treatment are registered by the surgeon immediately after surgery. The follow-up for revision will start on the day of primary days and last up to 90 days and 365 days after primary THA.

### Data sources

All outcome measures will be extracted from the following national and validated databases: *the Civil Registration System (CRS)*(*33*); *the DHR* (*28, 34, 35*); *the Danish National Patient Registry (DNRP)* (*36*); *The Hospital Acquired Infections Database (HAIBA)* (*37*); *the Danish National Prescription Registry (NPR)*(*38*) and *Statistics Denmark*.

<u>The Danish Civil Registration System (Centrale Person Register, CPR):</u> All Danish residents and citizens are assigned a unique and permanent individual identification number (CPR number) at birth or upon immigration. The register contains continuously updated information on migration and vital status including date of death. The CPR number goes through all Danish registries and enables an unambiguous linkage between registries and complete individual-level follow-up over time (33).

<u>The Danish National Patient Register (DNRP):</u> Contains data on all admissions and discharges from somatic hospitals in Denmark, including dates of admissions and discharges, surgical procedures performed, and up to twenty diagnoses for every discharge classified by the International Classification of Diseases (ICD) (39). The data includes outpatient and inpatient contacts and is linked to the patient’s CPR number (36).

<u>The Danish Hip Arthroplasty Register (DHR):</u> Collects data including type and duration of antibiotic treatment for all primary THAs and revisions performed in Denmark (34). It is mandatory for all surgeons performing THAs to report to the registry resulting in a high degree of completeness, with an overall completeness rate of 94% (35). The data on PJI has previously been validated (28). Furthermore, preoperative data include hospital code and laterality of the affected hip, date of surgery; thromboembolic prophylaxis; type of anesthesia; duration of surgery; type of acetabular and femoral component and their fixation and type, size, and material of the prosthetic femoral head and the acetabular liner (1). For revisions, the following are registered: Indication, prosthetic status before revision, the extent of revision, and the number of earlier revisions (40). The revisions performed within 90 and 365 days are located in the DHR. DHR is compared with DNRP for revisions and to determine the completeness of data.

<u>The Hospital Acquired Infections Database (HAIBA):</u> the database is an automated system for the surveillance of hospital-acquired infections. HAIBA monitors specific types of infections, using algorithms, which combine data from the DNPR and The Danish Microbiology Database (MiBa) 3aasasa. MiBa automatically collects all microbiology results from all departments of clinical microbiology in Denmark, since 2010. The data is stored electronically using the patient’s CPR number as the patient identifier (41). HAIBA provides continuous surveillance data, allowing for trend analysis (37).

<u>The Danish National Prescription Registry (NPR</u>) has recorded detailed information on prescriptions redeemed in Denmark since 1995. The NPR receives data recorded in the electronic dispensing systems of community pharmacies. The registry contains information related to the user, the prescriber, the dispensing pharmacy, and the drug prescribed and has previously been validated (38).

<u>Statistics Denmark</u> is a collection of registered data containing detailed information on the socioeconomic characteristics of all Danish citizens at the individual level. Information regarding family annual household income and liquid assets will be retrieved from The Income Statistics Register (42) containing more than 160 variables including salary savings of people with a Danish income. The data are primarily supplied by tax authorities. The Population Education Register (48) obtains information on the highest completed level of education and consists of data generated from administrative records of educational institutions and surveys. The Register-based Labour Force Statistics (RAS) obtains a description of the affiliation with the labor market. The registers are updated yearly and administered by the Danish government. Each individual is identified with an unambiguous CPR number (47).

### Outcomes

#### Primary Outcome

##### Risk of revision due to infection 90 days from index surgery

For the definition of infection, the definition of PJI is applied. Revision surgery is defined as a new surgical intervention for the first time after the primary THA including debridement alone or in combination with complete or partial removal or exchange of any implants.

The primary outcome is captured within 90 days of index surgery. For this trial, an initial 90-day surveillance period has been chosen as recommended by the National Healthcare Safety Network (NHSN 2022) (43) and studies confirm, that most infections following arthroplasty occur within the first 90 days after surgery (27, 44).

PJI is considered present when at least one of the following three criteria exists:

1. Two or more intraoperative deep-tissue samples of phenotypically indistinguishable bacteria isolated from at least three deep-tissue samples (45)
2. A PJI when an indication of deep infection is reported to DHR by the surgeon upon revision surgery (28)

The definition of PJI is based on a modified version of the definition provided by EBJIS (45), an International Consensus (15), and an algorithm developed to capture cases with PJI using national databases (27). For this trial, the definition of PJI is modified to include the most widely accepted definition of PJI with the main importance set to intraoperative cultures (2, 46). The definition has been modified to allow for the capture of PJI through databases and registries without review of medical files and the modifications are expected only to give minor non-significant changes for the capture of PJI (27, 28). Data will be extracted from DNRP, DHR, and HAIBA. Positive culture samples (aspirations, tissue biopsies, or fluid) must be obtained from the relevant hip joint.

Sinus tract communication with the joint or prosthesis visualization is expected to be captured as an indication of deep infection reported to DHR by the surgeon upon revision.

In contrast to EBJIS (45), we have not included histological examination of intraoperative tissue biopsies, erythrocyte sedimentation rate, white blood cell count, or biomarker analysis in joint fluid as these analyses are not routinely performed in Denmark. CRP levels must be interpreted with caution and cannot stand alone and are not included in our definition (27).

#### Secondary outcomes

The secondary outcomes are captured within 90 days of primary THA:

##### 1) Any revision after THA

Revision surgery is defined previously (see primary outcome). The rate of revision is defined as revision due to any cause within one year from primary THA surgery. The rate of revision will be recorded from DHR and DNPR.

##### 2) Potential PJI referred to as *PJI-likely*

Incidence of potential PJI. PJI-likely is defined as at least one of the two criteria being fulfilled:

1. One single intraoperatively obtained positive culture obtained from reoperation (aspiration fluid OR tissue biopsy) regardless of microorganism

These definitions of PJI-likely are based on a modified version of EBJIS (45) as described previously (see primary outcome) and the study by Milandt *et al*. (47) where first-time revisions with one positive culture were found to have a higher risk of re-revision for PJI.

Cases of PJI-likely will be captured in HAIBA and MIBA, and registration of antibiotic prescriptions in NPR. Positive culture samples (aspirations, tissue biopsies, or fluid) must be obtained from the relevant hip joint.

##### 3) Hospital-treated infections (Not PJI or PJI-likely)

Any Hospital-treated infection is defined as any first-time hospital admission with a primary or secondary infection diagnosis after discharge from index THA surgery. Hospital-treated infections are identified from DNPR based on ICD-10 codes listed in Appendix C. The list of infections includes chronic and any rare infections, to detect possible flare-ups in any possible ongoing infections.

##### 4) Community-based antibiotic use

The proportion of patients with at least one dispensing of any antibiotic after discharge from primary THA surgery. Community-based antibiotic use is a surrogate measure of any community-treated infection and is defined as at least one dispensing of narrow- and broad-spectrum antibiotics based on the Anatomical Therapeutic Chemical classification (ATC) codes. Medications are coded according to the ATC codes listed in Appendix C. All antibiotics in Denmark require prescriptions from a physician and these will be identified using NPR (38).

##### 5) Mortality

Mortality rate is defined by the date of death due to any cause within one year after primary THA surgery. Data will be collected from CRS.

##### 1) Risk of revision due to infection 365 days from index surgery

The secondary outcome measures the risk of revision due to infection following THA using the same definition of PJI as the primary outcome. The assessment period is extended up to 365 days post-index surgery, beyond the initial 90 days, to comprehensively capture both late and chronic PJIs.

### Data management

Eligible patients are identified from the Danish Hip Arthroplasty Registry and uploaded to the secure online research platform managed by Statistics Denmark, enabling linkage with data from the PCR, DNRP, NPR and HAIBA. All subsequent data handling and analyses will be conducted within this secure remote-access environment, accessible exclusively via the corresponding author’s digital ID. Data management and statistical analyses will be performed using R statistical software (48).

### Sample Size and Power Considerations

This study leverages a nationwide cohort of approximately 110,000 patients who underwent THA for osteoarthritis or secondary causes of osteoarthritis in Denmark between January 1, 2010, and December 31, 2020. It is expected that more than half of the patients received a prophylactic dosage exceeding a single dose. Because the sample size is defined by the national flow of patients in routine care, we did not conduct formal power or sample size calculations. The incidence of revision due to PJI in Denmark The PJI rate in Denmark has been reported to range from 0.5% to 5.1%, varying at department level, with a national average of 1.2% (95% CI: 1.0–1.4%) (49). To assess the potential for clinically meaningful harm, we adopted a non-inferiority framework. Based on a previous ongoing studies, we defined a non-inferiority margin of 0.1 percentage points (i.e., an absolute risk difference of 1 additional PJI per 1,000 patients), which we consider acceptable from a clinical standpoint (50).Given the large cohort size and expected distribution across exposure groups, the study should have sufficient precision to detect small differences in infection risk. For example, assuming similar event rates between groups and no clustering adjustment, the 95% confidence interval for the odds ratio is expected to be narrow, likely well within the defined non-inferiority margin. This allows for robust estimation of differences in infection risk, even with conservative model assumptions.

### Statistical methods

Descriptive statistics: Continuous variables will be summarized using means and standard deviations or medians and interquartile ranges (IQRs), as appropriate. Categorical variables will be reported as counts and percentages. Differences between treatment groups at baseline will be assessed using standardized differences (51); standardized differences are preferred over P-values because they provide a standardized, interpretable, and meaningful measure for assessing and addressing imbalances between groups in baseline characteristics, which is crucial for ensuring the validity of subsequent analyses.

To address potential treatment selection bias and confounding, propensity score methods will be used to emulate random assignment (30, 52). Comparative analyses will be conducted using three approaches: (1) unadjusted analyses without controlling for confounders, (2) stabilized inverse probability of treatment weighting (sIPTW) as the main method for de-confounding, and finally (3) propensity score matching (PSM) as a sensitivity analysis to assess the robustness of the findings. Propensity scores will be generated and applied as weights (sIPTW) to balance key confounding variables between the two treatment groups. Covariate balance before and after weighting will be assessed using standardized differences (SMDs), with an SMD <0.1 indicating acceptable balance (53, 54).

All statistical endpoints are binary and will be analyzed using weighted logistic regression models incorporating sIPTW. Results will be presented for each group as well the as the contrast between groups based on the odds ratio (and 95% confidence interval) derived from model coefficients, along with absolute and relative risk differences and their corresponding 95% confidence intervals, calculated using average marginal effects. Confidence intervals will be estimated using standard errors.

The main analyses will be adjusted for calendar time, and known risk factors, as outlined in Appendix B. Hospital will be included as a random effect to account for clustering at center level. Propensity scores will be estimated using a logistic mixed-effects model including the following covariates (52):

- Year of surgery (continuous variable), included as a fixed effect
- Comorbidity status (Elixhauser: <0, 0, 1-4, ≥5) (categorical variable), included as a fixed effect
- Age (continuous variable), included as a fixed effect
- Sex (binary variable), included as a fixed effect
- Education level (categorical variable), included as a fixed effect
- Living alone or co-living (binary variable), included as a fixed effect
- Income (categorical variable), included as a fixed effect
- Primary diagnosis (categorical variable), included as a fixed effect
- Previous hip surgery (binary variable), included as a fixed effect
- Type of fixation (cemented, uncemented, hybrid) (categorical variable), included as a fixed effect
- Type of cement (antibiotic-loaded cement, plain cement, no cement)
- Duration of surgery (continuous variable), included as a fixed effect

A separate analysis will be conducted for the cohort from 2017 to 2020 Here the following variables will be included too:

- BMI (categorical variable ≤30, >30), included as a fixed effect
- Morbid obesity (binary variable yes/no, BMI ≥40), included as a fixed effect

Each unit will be assigned a weight based on the inverse probability of receiving the treatment they actually receive. Specifically, patients in the treatment group will be weighted by the inverse of their propensity score (1/PS), and control patients by the inverse of one minus their propensity score [1/(1–PS)]. To reduce variance and improve covariate balance, these weights will be stabilized by multiplying each unit’s weight by the marginal probability of being in their respective treatment group (55). Patients in the treatment group with low propensity scores and controls with high propensity scores will therefore be given more weight in the analysis.

Pre-specified subgroup analyses will be performed as outlined in Appendix E. Furthermore, an extra model will be performed for the sensitivity analyses with supplementary adjustment for BMI to study the effect of this missing variables.

### Loss to follow-up and Missing Data

Generally, no data imputation will be performed. Missing data on potential confounders will be assumed missing completely at random. Patients who die or emigrate within 90 days following surgery will be excluded from outcome analyses. Given the low 90-day mortality rate after THA, we do not expect this exclusion to substantially impact the study results. Consequently, the main analyses will be based on the data ‘As Observed’ which is valid assuming that data is ‘Missing Completely At Random’ (MCAR) (51). For sensitivity analysis, we will also conduct the primary analysis using both worst-case and best-case imputation to assess the robustness of the main findings (56).

## ETHICAL ASPECTS

The project is registered in accordance with Danish legislation. No additional ethical approval is required. All data will be handled confidentially and in compliance with Danish data protection regulations. The study protocol will be uploaded to www.clinicaltrials.gov prior to conduction to ensure transparency. All data used in this study are stored securely on the servers of Statistics Denmark. In accordance with Statistics Denmark’s data protection policies, the data cannot be shared in raw or anonymized form. However, researchers may apply for access to the same underlying data directly through Statistics Denmark.

## PERSPECTIVES

The use of antibiotics is regarded as the primary driver of worldwide antimicrobial resistance that may pose a threat to global health (17–19, 57). Therefore, there is an urgent need to establish evidence to potentially change clinical practice on antibiotic prophylaxis dosages in the future. Previous studies are based on data from national registries. The registration of infection is reported to DHR immediately after revision surgery. Therefore, microbiology results are not available at the time of registration and hence not taken into consideration when the cause of revision is reported. This planned study applies an advanced approach that allows for a more accurate capture of the PJI incidence. This may add valuable knowledge on the effect of different antibiotic duration practices on the risk of PJI.

## COLLABORATORS/PROJECT TEAM

<u>Primary investigator:</u> Armita Armina Abedi, MD, PhD student

<u>Secondary investigator:</u> Marie Anneberg Brahe, Department of Clinical Epidemiology, Aarhus University Hospital, Aarhus, Denmark.

<u>Main Supervisor:</u> Søren Overgaard: MD, DMSc, Professor, Head of Research, Department of Orthopedic surgery, Bispebjerg and Frederiksberg Hospital, Denmark.

<u>Secondary Supervisors:</u>

Alma Becic Pedersen, MD, PhD, DMsc, Professor, Department of Clinical Epidemiology, Aarhus University Hospital, Aarhus, Denmark.

Claus Varnum, MD, PhD, Associate Professor, Department of Orthopedics, Lillebaelt Hospital - Vejle, Vejle, Denmark.

Robin Christensen, BSc, MSc, PhD, Professor of Biostatistics and Clinical Epidemiology, Section for Biostatistics and Evidence-Based Research, the Parker Institute, Bispebjerg and Frederiksberg Hospital, Copenhagen & Department of Clinical Research, University of Southern Denmark, Odense University Hospital, Denmark.

Database manager: Armita Armina Abedi and Marie Anneberg Brahe

Senior Statistician: Robin Christensen

The project is approved by the Department of Orthopedic Surgery and Traumatology, Bispebjerg, Denmark and there is a designated working place at Bispebjerg Hospital.

## Data Availability

The data presented in this study is stored on the servers of Statistics Denmark. Due to the rules for data protection at Statistics Denmark, it is not possible to share the data, neither in raw nor in anonymized form. However, other researchers can apply for permission to access the same raw data at Statistics Denmark.

## APPENDICES

## Appendix A

**Table 1.**
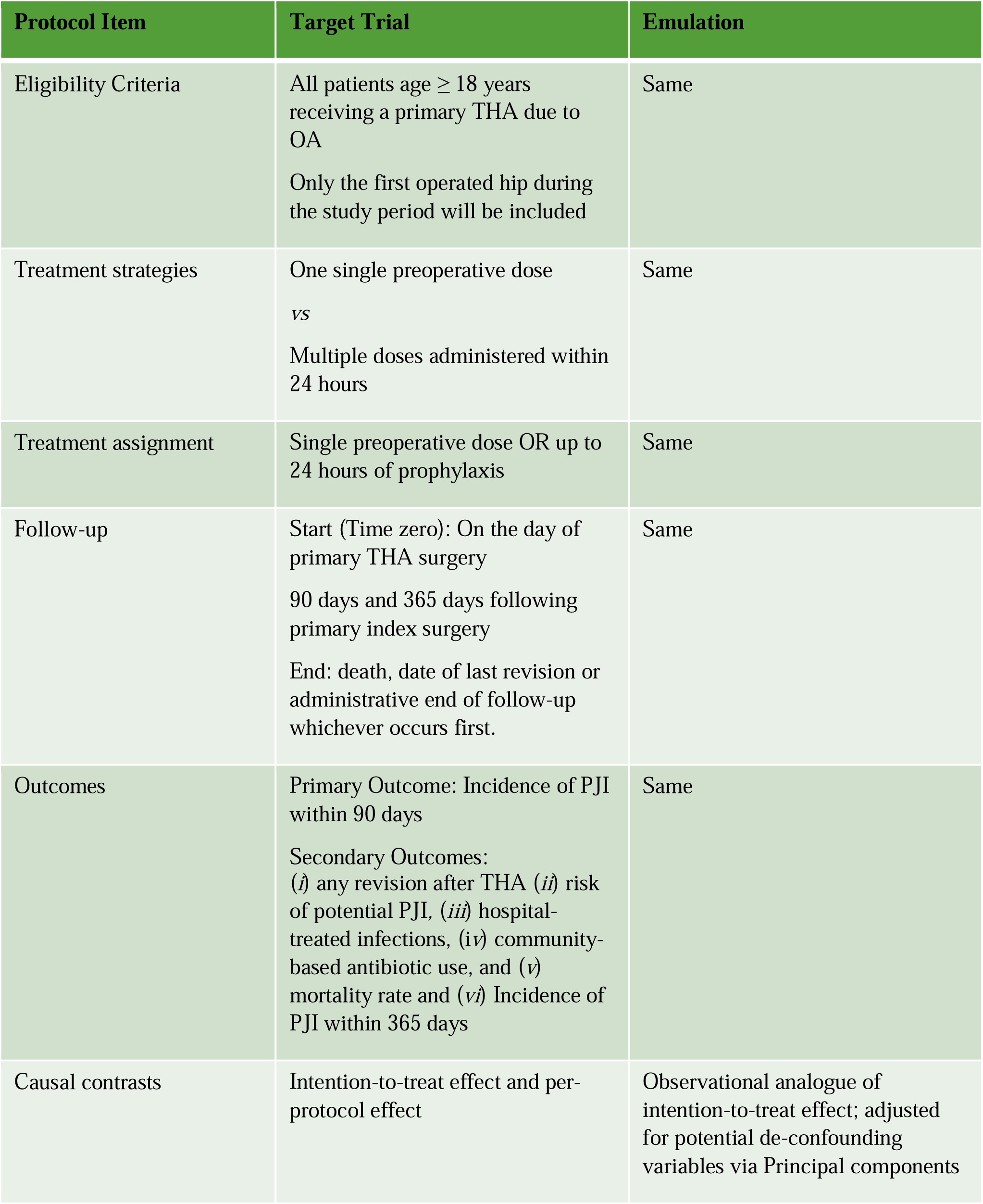
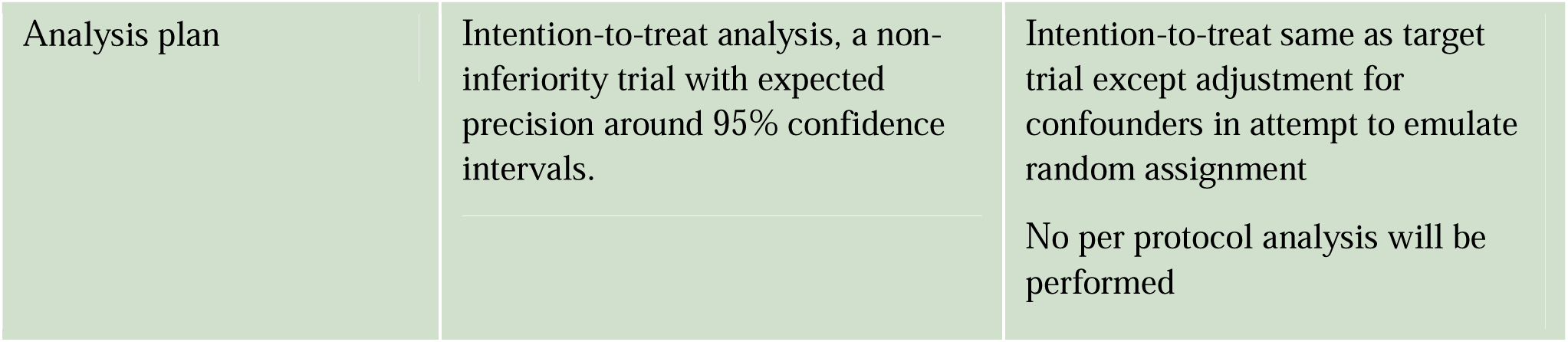
Specification and emulation of the target trial

## Appendix B

Direct acyclic graph used for choosing the variables to adjust for

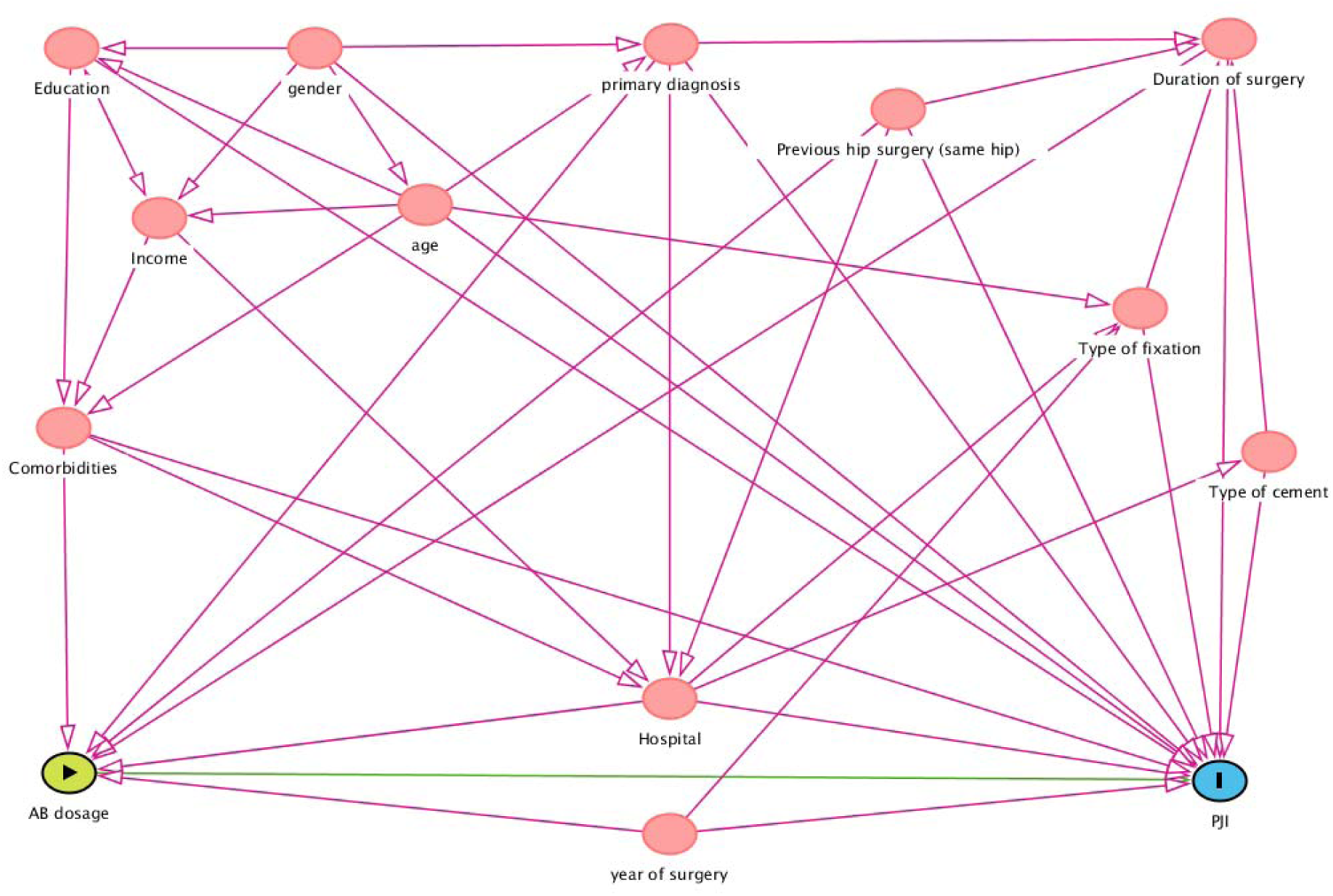

## Appendix C

International Classification of Diseases, Tenth Revision (ICD-10) codes used to identify hospital-treated infection.

Codes used to identify any hospital-treated infection:
| <u>Infections</u> | <u>ICD-10 diagnosis codes</u> |
| --- | --- |
| Any hospital-treated (inpatient or outpatient) infection | A20 -A38, A42 -A44, A48 -A49, A65 -A79, A3, A49.9, A39.4, A40 -A41, B37.7, A32.7, A54.8G, A02.1, A22.7, A26.7, A42.7, A28.2B, A06.5, A54.1, B43, D73.3, E06.0A, E23.6A, E32.1, G06, G07, H00.0A, H05.0A, H44.0A, H60.0, J34.0A, J36, J38.3D, J38.7G, J39.0, J39.1, J39.8A, J85.1, J85.2, J85.3, K04.6, K04.7, K11.3, K12.2, K13.0A, K14.0A, K20.9A, K35.3A, K35.3B, K57.0, K57.2, K57.4, K57.8, K61, K63.0, K65.0, K75.0, K81.0A, K85.8A, L02, L05.0, L05.9, M60.8A, M86.8A, M86.9A, N15.1, N34.0, N41.2, N45.0, N48.2, N49.2A, N61.9A, N61.9B, N70.0A, N70.0B, N71.0A, N73.0A, N73.0B, N73.2A, N73.2B, N73.3A, N73.5A, N73.8A, N73.8C, N75.1, N76.4, N76.8A, **Except:** A54.1B, B43.0, B43.8, B43.9, K57.0B, K57.0C, K57.2B, K57.2C, K57.4A, K65.0M, K65.0N, K65.0O, K65.0P, A46, H01.0, H03, H60.0, H60.1, H60.2, H60.3, H62, K12.2, K13.0, K61, M72.6, L01, L08, L03, J34.0, L00, L02, L04, L05, L06, L07, L30.3, L73.8, H00, H01.0, H03.0, H03.1, H04.3, H05.0, H06.1, H10, H13.0, H13.1, H15.0, H19.1, H19.2, H22.0, H32.0, H44.0, H44.1, H60, H61.0, H62.0, H62.1, H62.2, H62.3, H65, H66, H67.0, H67.1, H68, H70, H73.0, H75.0, H83.0, H94.0 **Except:** H60.4, H60.4A, H60.5, H60.5B, H60.8, H60.8A, H65.2, H65.3, H65.4, H65.4C, H66.1, H66.2, H66.3, H68.1, H70.1, H70.8, G00 -07, A80 -A89, G00, G01, G02, G03, A32.1, A39.0, A17.0, A20.3, A87, A54.8D, A02.2C, B37.5, B00.3, B01.0, B02.1, B05.1, B26.1, B38.4, A00 -A09, K35, K37, K57.0, K57.2, K57.4, K57.8, K61, K63.0, K65.0, K65.9, K67, K75.0, K75.1, K80.0, K80.3, K80.4, K81.0, K81.9, K83.0, K85.9, I00 -I02, I30.1, I32.0, I33, I38, I40.0, I39.8, B37.6, J00 -J06, J36, J39.0, J39.1, J12 -J18, J20 -J22, J44.0, J85.1, J86, J20 -J22, J34.0, J35.0, J38.3C, J38.3D, J38.7B, J38.7F, J38.7G, **Except:** J34.0E, J34.0F, J34.0G, J34.0H, N10, N11, N12, N15.1, N15.9, N30, N33.0, N34, N39.0, N08.0, N13.6, N16.0, N28.8D, N28.8E, N28.8F, N29.0, N29.1, **Except:** N30.1, N30.2, N30.4, A50 -A64, N41, N45, N48.1, N48.2, N49, N51.1, N51.2, N70 -77, O23, O26.4, O41.1, O74.0, O75.3, O85, O86, O88.3, O91, O98, M00, M01, M86, M63.0, M63.2, T80.2, T81.4, T82.6, T82.7, T83.5, T83.6, T84.5, T84.6, T84.7, T85.7, T88.0, T89.9, B90 -B99, K04.0, K05.2 |

Codes for specific hospital-treated infections:
| <u>Infections</u> | <u>ICD-10 diagnosis codes</u> |
| --- | --- |
| Pneumonia | J12-J18 |
| Urinary tract infections | N10, N11, N12, N15.1, N15.9, N30, N33.0, N34, N39.0, N08.0, N13.6, N16.0, N28.8D, N28.8E, N28.8F, N29.0, N29.1, Except: N30.1, N30.2, N30.4 |

## Appendix D

Anatomical Therapeutical Chemical Classification System (ATC) codes used to identify community-based antibiotic use.

Appendix D Anatomical Therapeutical Chemical Classification System (ATC) codes used to identify community-based antibiotic use.
| <u>Narrow spectrum antibiotics</u> | <u>ATC-code</u> |
| --- | --- |
| Beta-lactamase sensitive penicillins, Beta-lactamase resistant penicillins, first-generation cephalosporins. | J01CE, J01CF, J01DB |
| <u>Broad-spectrum antibiotics</u> | <u>ATC-code</u> |
| Cephalosporins (second generation, third-generation, fourth-generation) Carbapenem, Other cephalosporins and penems, Combinations of penicillins, including beta-lactamase inhibitors, penicillins with extended spectrum, macrolides, lincosamides and streptogramins, trimethoprim and sulphonamides, Fluoroquinolones, Tetracyclines. | J01DC, J01DD, J01DE, J01DH, J01DI, J01CR, J01CA, J01F, J01E, J01MA, J01AA |

## APPENDIX E Subgroup Analysis

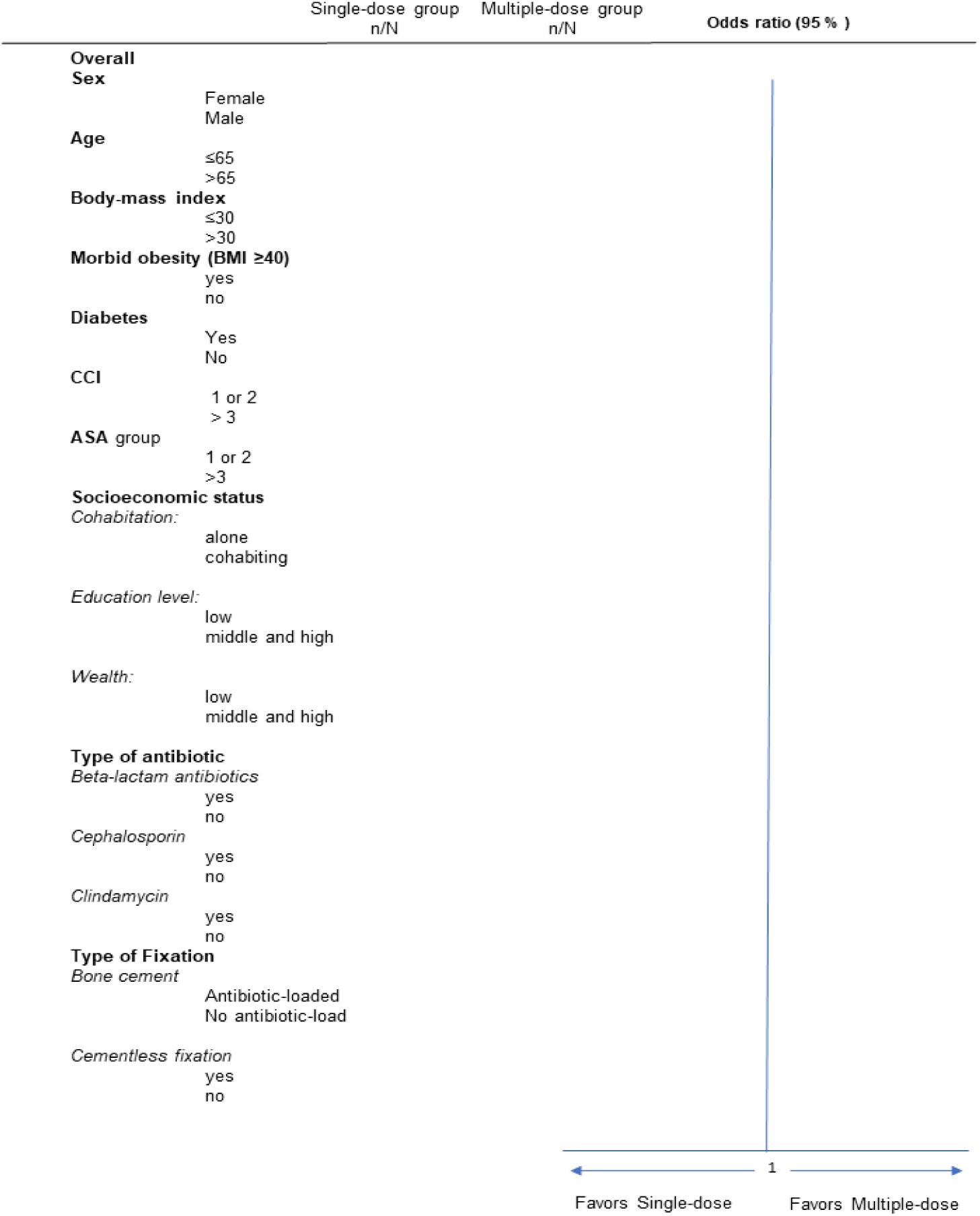

## References

1. Register D. DHR Årsrapport 2021. Annual reports The Danish Hip Arthorplasy Register. 2021. 2021 [Available from: http://danskhoftealloplastikregister.dk/wp-content/uploads/2022/07/DHR-aarsrapport-2021_Udgivet-2022_offentliggjort-version.pdf.

2. Zimmerli W, Trampuz A, Ochsner PE. Prosthetic-joint infections. N Engl J Med. 2004;351(16):1645–54.

3. Zmistowski B, Karam JA, Durinka JB, Casper DS, Parvizi J. Periprosthetic joint infection increases the risk of one-year mortality. J Bone Joint Surg Am. 2013;95(24):2177–84.

4. Bozic KJ, Ries MD. The impact of infection after total hip arthroplasty on hospital and surgeon resource utilization. J Bone Joint Surg Am. 2005;87(8):1746–51.

5. Gundtoft PH, Pedersen AB, Varnum C, Overgaard S. Increased Mortality After Prosthetic Joint Infection in Primary THA. Clin Orthop Relat Res. 2017;475(11):2623–31.

6. Carlsson AK, Lidgren L, Lindberg L. Prophylactic antibiotics against early and late deep infections after total hip replacements. Acta Orthop Scand. 1977;48(4):405–10.

7. Doyon F, Evrard J, Mazas F, Hill C. Long-term results of prophylactic cefazolin versus placebo in total hip replacement. Lancet. 1987;1(8537):860.

8. Hill C, Flamant R, Mazas F, Evrard J. Prophylactic cefazolin versus placebo in total hip replacement. Report of a multicentre double-blind randomised trial. Lancet. 1981;1(8224):795–6.

9. van Kasteren ME, Manniën J, Ott A, Kullberg BJ, de Boer AS, Gyssens IC. Antibiotic prophylaxis and the risk of surgical site infections following total hip arthroplasty: timely administration is the most important factor. Clin Infect Dis. 2007;44(7):921–7.

10. AlBuhairan B, Hind D, Hutchinson A. Antibiotic prophylaxis for wound infections in total joint arthroplasty: a systematic review. J Bone Joint Surg Br. 2008;90(7):915–9.

11. Berrios-Torres SI, Umscheid CA, Bratzler DW, Leas B, Stone EC, Kelz RR, et al. Centers for Disease Control and Prevention Guideline for the Prevention of Surgical Site Infection, 2017. JAMA Surg. 2017;152(8):784–91.

12. World Health O. Global guidelines for the prevention of surgical site infection. 2nd ed. ed. Geneva: World Health Organization; 2018 2018.

13. Bratzler DW, Dellinger EP, Olsen KM, Perl TM, Auwaerter PG, Bolon MK, et al. Clinical practice guidelines for antimicrobial prophylaxis in surgery. Am J Health Syst Pharm. 2013;70(3):195–283.

14. Hansen E, Belden K, Silibovsky R, Vogt M, Arnold W, Bicanic G, et al. Perioperative antibiotics. J Orthop Res. 2014;32 Suppl 1:S31–59.

15. Parvizi J, Gehrke T Fau - Chen AF, Chen AF. Proceedings of the International Consensus on Periprosthetic Joint Infection. (2049–4408 (Electronic)).

16. Knæ-alloplastikkirurgi DSfHo. Kort Klinisk Retningslinje, Ortopædisk Kirurgi. Antibiotika profylakse ved alloplastikkirurgi hofte/knæ. 2017:5.

17. Centers for Disease C, Prevention. Vital signs: carbapenem-resistant Enterobacteriaceae. MMWR Morb Mortal Wkly Rep. 2013;62(9):165–70.

18. Jacoby GA, Munoz-Price LS. The new beta-lactamases. N Engl J Med. 2005;352(4):380–91.

19. Laxminarayan R, Duse A, Wattal C, Zaidi AK, Wertheim HF, Sumpradit N, et al. Antibiotic resistance-the need for global solutions. Lancet Infect Dis. 2013;13(12):1057–98.

20. Thornley P, Evaniew N, Riediger M, Winemaker M, Bhandari M, Ghert M. Postoperative antibiotic prophylaxis in total hip and knee arthroplasty: a systematic review and meta-analysis of randomized controlled trials. CMAJ Open. 2015;3(3):E338–43.

21. Siddiqi A, Forte SA, Docter S, Bryant D, Sheth NP, Chen AF. Perioperative Antibiotic Prophylaxis in Total Joint Arthroplasty: A Systematic Review and Meta-Analysis. J Bone Joint Surg Am. 2019;101(9):828–42.

22. Voigt J, Mosier M, Darouiche R. Systematic review and meta-analysis of randomized controlled trials of antibiotics and antiseptics for preventing infection in people receiving primary total hip and knee prostheses. Antimicrob Agents Chemother. 2015;59(11):6696–707.

23. Tan TL, Shohat N, Rondon AJ, Foltz C, Goswami K, Ryan SP, et al. Perioperative Antibiotic Prophylaxis in Total Joint Arthroplasty: A Single Dose Is as Effective as Multiple Doses. J Bone Joint Surg Am. 2019;101(5):429–37.

24. Veltman ES, Lenguerrand E, Moojen DJF, Whitehouse MR, Nelissen R, Blom AW, et al. Similar risk of complete revision for infection with single-dose versus multiple-dose antibiotic prophylaxis in primary arthroplasty of the hip and knee: results of an observational cohort study in the Dutch Arthroplasty Register in 242,179 patients. Acta Orthop. 2020;91(6):794–800.

25. Veltman ES, Moojen DJF, Nelissen RG, Poolman RW. Antibiotic Prophylaxis and DAIR Treatment in Primary Total Hip and Knee Arthroplasty, A National Survey in The Netherlands. (2206-3552 (Print)).

26. Lindgren JV, Gordon M Fau - Wretenberg P, Wretenberg P Fau - Kärrholm J, Kärrholm J Fau - Garellick G, Garellick G. Validation of reoperations due to infection in the Swedish Hip Arthroplasty Register. (1471–2474 (Electronic)).

27. Gundtoft PH, Overgaard S, Schonheyder HC, Moller JK, Kjaersgaard-Andersen P, Pedersen AB. The “true” incidence of surgically treated deep prosthetic joint infection after 32,896 primary total hip arthroplasties: a prospective cohort study. Acta Orthop. 2015;86(3):326–34.

28. Gundtoft PH, Pedersen AB, Schonheyder HC, Overgaard S. Validation of the diagnosis ‘prosthetic joint infection’ in the Danish Hip Arthroplasty Register. Bone Joint J. 2016;98-B(3):320–5.

29. Hernán Ma Fau - Robins JM, Robins JM. Using Big Data to Emulate a Target Trial When a Randomized Trial Is Not Available. (1476–6256 (Electronic)).

30. Matthews AA, Danaei G, Islam N, Kurth T. Target trial emulation: applying principles of randomised trials to observational studies. (1756–1833 (Electronic)).

31. Hernán MA, Wang W, Leaf DE. Target Trial Emulation: A Framework for Causal Inference From Observational Data. (1538–3598 (Electronic)).

32. Hernán MA RJ. Causal Inference: What If: Boca Raton: Chapman & Hall/CRC; 2020, Revised 2023.

33. Schmidt M, Pedersen L, Sørensen HT. The Danish Civil Registration System as a tool in epidemiology. European Journal of Epidemiology. 2014;29(8):541–9.

34. Gundtoft PH, Varnum C, Pedersen AB, Overgaard S. The Danish Hip Arthroplasty Register. Clin Epidemiol. 2016;8:509–14.

35. Pedersen A, Johnsen S, Overgaard S, Søballe K, Sørensen HT, Lucht U. Registration in the danish hip arthroplasty registry: completeness of total hip arthroplasties and positive predictive value of registered diagnosis and postoperative complications. Acta Orthop Scand. 2004;75(4):434–41.

36. Schmidt M, Schmidt SA, Sandegaard JL, Ehrenstein V, Pedersen L, Sørensen HT. The Danish National Patient Registry: a review of content, data quality, and research potential. Clin Epidemiol. 2015;7:449–90.

37. (SSI) SSI. HAIBA – Hospital-Acquired Infections database 2015 [updated 4th March 2015. Available from: https://en.ssi.dk/news/epi-news/2015/no-9---2015.

38. Pottegard A, Schmidt SAJ, Wallach-Kildemoes H, Sorensen HT, Hallas J, Schmidt M. Data Resource Profile: The Danish National Prescription Registry. Int J Epidemiol. 2017;46(3):798-f.

39. WHO International Classification of Diseases. 2023 Jan 2023 [Available from: https://www.who.int/standards/classifications/classification-of-diseases.

40. Pedersen A, Johnsen S Fau - Overgaard S, Overgaard S Fau - Søballe K, Søballe K Fau - Sørensen HT, Sørensen Ht Fau - Lucht U, Lucht U. Registration in the danish hip arthroplasty registry: completeness of total hip arthroplasties and positive predictive value of registered diagnosis and postoperative complications. (0001–6470 (Print)).

41. Voldstedlund M, Haarh M, Molbak K, MiBa Board of R. The Danish Microbiology Database (MiBa) 2010 to 2013. Euro Surveill. 2014;19(1).

42. Baadsgaard M, Quitzau J. Danish registers on personal income and transfer payments. (1651–1905 (Electronic)).

43. (NHSN) NHSN. NHSN Surgical Site Infection Event (SSI) January 2022.pdf. 2022.

44. Yokoe DS, Avery TR, Platt R, Huang SS. Reporting surgical site infections following total hip and knee arthroplasty: impact of limiting surveillance to the operative hospital. Clin Infect Dis. 2013;57(9):1282–8.

45. McNally M, Sousa R, Wouthuyzen-Bakker M, Chen AF, Soriano A, Vogely HC, et al. Infographic: The EBJIS definition of periprosthetic joint infection. Bone Joint J. 2021;103-B(1):16–7.

46. Parvizi J, Gehrke T, Mont MA, Callaghan JJ. Introduction: Proceedings of International Consensus on Orthopedic Infections. The Journal of Arthroplasty. 2019;34(2):S1–S2.

47. Milandt NR, Gundtoft PH, Overgaard S. A Single Positive Tissue Culture Increases the Risk of Rerevision of Clinically Aseptic THA: A National Register Study. Clin Orthop Relat Res. 2019;477(6):1372–81.

48. R: A language and environment for statistical computing. Vienne, Autria: R Foundation for Statistical Computing. [Internet]. 2010. Available from: http://www.R-project.org.

49. Annual reports The Danish Hip Arthorplasy Register 2023. DHR-årsrapport-2023.2024. Available from: https://www.sundk.dk/media/ci5smyq5/90559b0251eb4824b60ca2b6fd98e4ca.pdf.

50. Abedi AA, Varnum C, Pedersen AB, Gromov K, Hallas J, Iversen P, et al. Effect of single versus multiple prophylactic antibiotic doses on prosthetic joint infections following primary total hip arthroplasty in patients with osteoarthritis at public and private hospitals in Denmark: protocol for a nationwide cross-over, cluster randomised, non-inferiority trial [The Pro-Hip-Quality Trial]. BMJ Open. 2023;13(8):e071487.

51. Christensen R, Ranstam J, Overgaard S, Wagner P. Guidelines for a structured manuscript: Statistical methods and reporting in biomedical research journals. Acta Orthop. 2023;94:243–9.

52. Rosenbaum PR, Rubin DB. The central role of the propensity score in observational studies for causal effects. Biometrika. 1983;70(1):41–55.

53. Austin PC. Using the Standardized Difference to Compare the Prevalence of a Binary Variable Between Two Groups in Observational Research. Communications in Statistics - Simulation and Computation. 2009;38:1228–34.

54. Austin PC, Stuart EA. Moving towards best practice when using inverse probability of treatment weighting (IPTW) using the propensity score to estimate causal treatment effects in observational studies. (1097–0258 (Electronic)).

55. Austin PC, Stuart EA. Moving towards best practice when using inverse probability of treatment weighting (IPTW) using the propensity score to estimate causal treatment effects in observational studies. Stat Med. 2015;34(28):3661–79.

56. Liublinska V, Rubin DB. Sensitivity analysis for a partially missing binary outcome in a two-arm randomized clinical trial. Stat Med. 2014;33(24):4170–85.

57. Månsson E, Bech Johannesen T, Nilsdotter-Augustinsson Å, Söderquist B, Stegger M. Comparative genomics of Staphylococcus epidermidis from prosthetic-joint infections and nares highlights genetic traits associated with antimicrobial resistance, not virulence. LID - 10.1099/mgen.0.000504 [doi] LID - 000504. (2057–5858 (Electronic)).

